# Harmonization of Later-Life Cognitive Function Across National Contexts: Results from the Harmonized Cognitive Assessment Protocols (HCAPs)

**DOI:** 10.1101/2023.06.09.23291217

**Authors:** Alden L. Gross, Chihua Li, Emily M. Briceno, Miguel Arce Rentería, Richard N. Jones, Kenneth M. Langa, Jennifer J. Manly, Emma L. Nichols, David Weir, Rebeca Wong, Lisa Berkman, Jinkook Lee, Lindsay C. Kobayashi

## Abstract

**Background:** The Harmonized Cognitive Assessment Protocol (HCAP) is an innovative instrument for cross-national comparisons of later-life cognitive function, yet its suitability across diverse populations is unknown. We aimed to harmonize general and domain-specific cognitive scores from HCAPs across six countries, and evaluate precision and criterion validity of the resulting harmonized scores.

**Methods:** We statistically harmonized general and domain-specific cognitive function across the six publicly available HCAP partner studies in the United States, England, India, Mexico, China, and South Africa (N=21,141). We used an item banking approach that leveraged common cognitive test items across studies and tests that were unique to studies, as identified by a multidisciplinary expert panel. We generated harmonized factor scores for general and domain-specific cognitive function using serially estimated graded-response item response theory (IRT) models. We evaluated precision of the factor scores using test information plots and criterion validity using age, gender, and educational attainment.

**Findings:** IRT models of cognitive function in each country fit well. We compared measurement reliability of the harmonized general cognitive function factor across each cohort using test information plots; marginal reliability was high (r> 0·90) for 93% of respondents across six countries. In each country, general cognitive function scores were lower with older ages and higher with greater levels of educational attainment.

**Interpretation:** We statistically harmonized cognitive function measures across six large, population-based studies of cognitive aging in the US, England, India, Mexico, China, and South Africa. Precision of the estimated scores was excellent. This work provides a foundation for international networks of researchers to make stronger inferences and direct comparisons of cross-national associations of risk factors for cognitive outcomes.

**Funding:** National Institute on Aging (R01 AG070953, R01 AG030153, R01 AG051125, U01 AG058499; U24 AG065182; R01AG051158)

## Introduction

Alzheimer’s disease and related dementias, for which cognitive decline is the hallmark symptom, are a major global public health, clinical, and policy challenge. Although much research on risk and protective factors for dementia has been conducted in high-income countries it is anticipated that three-quarters of the 152 million persons with dementia will be living in low- and middle-income countries in the coming decades.^1–3^ Differences in the distributions of potential risk factors and cultural and demographic factors that impact dementia across countries makes cross-national research imperative.

To facilitate cross-national comparisons of later-life cognitive outcomes, measurement instruments must validly measure cognitive function across populations with diverse cultural, educational, social, economic, and political contexts. To that end, the Harmonized Cognitive Assessment Protocol (HCAP) has been developed and implemented in International Partner Studies (IPS) of the US Health and Retirement Study (HRS).^4^ The HCAP network represents the largest concerted global effort to-date to conduct harmonized large-scale population-representative studies of cognitive aging and dementia.

Although the HCAP was designed collaboratively to ensure its comparability across countries, necessary adaptations were made to its individual test items, test administrations, and scoring procedures to accommodate different languages, cultures, and levels of literacy and numeracy of its respondents.^15^ The impacts of these adaptations on the performance, reliability, and validity of the HCAP cognitive test items are only beginning to be understood^15,20^, which may limit cross-national utility of the HCAP battery. The goal of this study was to conduct statistical harmonization of the HCAP instruments fielded in the United States, England, India, Mexico, China, and South Africa. Statistical harmonization involved assigning cognitive test items to domains, determining which test items were common and which were unique across countries, deriving harmonized factor scores for general and specific cognitive domains, and estimating the reliability and validity of the harmonized factor scores.

## Methods

### Participants

The Health and Retirement Study in the United States (HRS) and its International Partner Studies (IPS) are large, population-based studies of aging. Between 2016 and 2019, six such studies administered Harmonized Cognitive Assessment Protocols (HCAPs) to participants from each core IPS. They included the HRS, the English Longitudinal Study on Ageing (ELSA), the Longitudinal Aging Study in India (LASI), the Mexican Health and Aging Study (MHAS), the China Health and Retirement Longitudinal Study (CHARLS), and Health and Aging in Africa: A Longitudinal Study of an INDEPTH Community in South Africa (HAALSI). Details of HCAP administration in each cohort, eligibility, timing, and sample sizes are summarized in **Supplemental Table 1**.^4–9^ The HCAP aims to provide a detailed assessment of cognitive function of older adults that is flexible, yet comparable across populations in countries with diverse cultural, educational, social, economic, and political contexts. The HCAP network ultimately intends to provide comparable estimates of dementia and mild cognitive impairment prevalence across countries, and to exploit cross-national variation in key risk and protective factors to better understand the determinants of later-life cognition, cognitive aging, and dementia.^61^

The HCAPs in the US and Mexico randomly sampled participants from the core studies who did not need a proxy interview in the previous core interview wave, and HRS-HCAP further included a random sampling of N=219 participants interviewed by proxy in the 2016 HRS core wave.^4,7^ To ensure adequate sample sizes of participants with dementia, HCAPs in England, India, and South Africa recruited participants with low cognitive function.^5,8,9^ All parent studies were nationally representative, with the exception of HAALSI in South Africa, which is a representative sample from the Agincourt sub-district in northeastern South Africa.^10^ All participants consented to research and IRBs at local institutions approved each IPS and its respective HCAP.

### Variables

Cognitive test battery. Details of the original battery of 17 cognitive tests in HCAP are available in Langa et al.^4^ By design, each HCAP study administered as close to the same battery of tests as was feasible. We granularized these batteries to 30-51 cognitive test indicators in each HCAP, as shown in Figure 1 and Supplemental Table 2.

**Figure 1.**
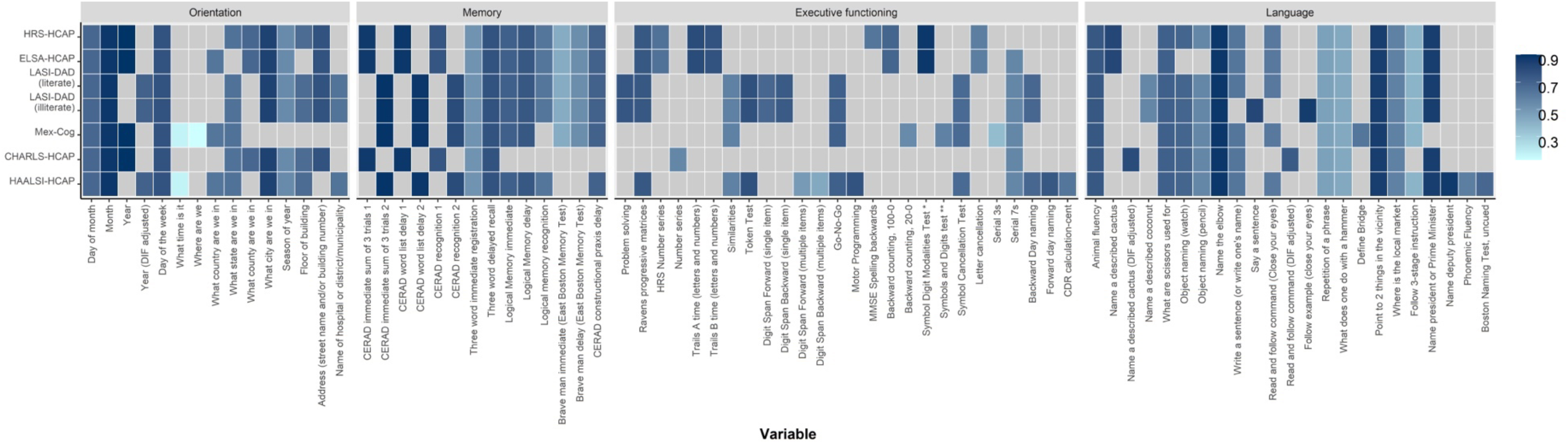
Heatmap of cognitive test items and their overlap across each study: Results from HCAP studies (N=21,141)

Each cognitive test item was assigned to a domain based on *a priori* theory, combined with empirical analyses demonstrating which test items fit well into a domain using factor analysis methods.^11–13^ Assigning test items to domains is essential to statistical harmonization, also referred to as co-calibration, as this process relies on the presence of equivalent or comparable cognitive test items across one or more studies. If cognitive test items are presumed to be the same across HCAP studies, but are in fact different (e.g., a different test; the same test with different stimuli, administration, or scoring procedures), such methodological differences could contribute to artifactual differences in the observed cognitive scores between studies. These artifactual differences would imperil the quality of cross-national inferences drawn from the derived summary cognitive scores.

To determine the comparability of cognitive test items across HCAPs, we convened an expert panel of neuropsychologists, epidemiologists, persons with cultural/linguistic expertise, and psychometricians with working knowledge of cross-cultural neuropsychology and administration of the HCAPs to conduct pre-statistical harmonization of cognitive test items. This group used available materials including codebooks, interviewer training manuals, and personal communication with study investigators and coordinators to document differences in test item content and administration across HCAPs and to determine which differences were substantial and whether cultural or language demands differed for each test. Considerations made for each cognitive test item have been described previously.^14,15^ Using the HRS HCAP as the reference, two neuropsychologists rated items from all other HCAPs as a confident linking item that is very likely to be comparable, a tentative linking item, or a non-linking item based on available information.^15^

### Covariates

Age, gender, and highest educational attainment were collected in core IPS interviews. We scaled educational attainment in each country to the 2011 International Standard Classification of Education.^16^

### Analysis plan

#### Descriptive analyses

We described demographic characteristics and cognitive tests using means with standard deviations and counts with percentages. We identified overlapping and unique cognitive test items by HCAP.

#### HCAP-specific factor analyses

We estimated confirmatory factor analysis (CFA) models for cognitive domains of general cognitive function, memory, executive function, orientation, and language separately in each HCAP study, without regard to items in common across studies. The goal of this series of psychometric models was to illustrate that similar organizations of cognitive test items fit well across countries.^12,17^ We ascertained model fit using three standard absolute fit statistics: the Root Mean Square Error of Approximation (RMSEA), Comparative Fit Index (CFI), and Standardized Root Mean Residual (SRMR).^18^ When possible, we attempted to improve model fit through the use of bifactor models to address additional correlations between theoretically similar items (e.g., Trail Making Test, parts A and B, or immediate and delayed recall).^11,19^ Using the combination of these three fit statistics, we characterized model fit as perfect, good, adequate, or poor, using previously described rubric.^12^

#### Statistical harmonization via item banking

Following the estimation of CFAs within each HCAP study, we statistically harmonized scores for each cognitive domain across countries using an item banking approach.^20^ A flowchart in **Supplemental Figure 1** illustrates this approach. For each cognitive domain, we serially estimated CFAs in each study, sequentially fixing model parameters for items determined to be comparable to those in previous studies to their corresponding values from previous studies. The order of studies was HRS-HCAP, ELSA-HCAP, LASI-DAD, Mex-Cog, CHARLS-HCAP, and HAALSI-HCAP. LASI-DAD was split into literate (N=1777, 43%) and illiterate (N=2139, 57%) subgroups due to administration differences in some tests. Because there is no natural scaling in latent variable space, the mean and variance of the factor score (general cognitive function, memory, executive function, orientation, or language) were set to 0 and 1, respectively, beginning with the HRS-HCAP as the reference. The factor score is estimated based on all of the items in the CFA. The CFA models estimated two relevant parameters for each cognitive test item: factor loadings and item thresholds (for categorical items) or intercepts (for continuous items). Factor loadings characterize how strongly correlated a given cognitive test item is with the other cognitive test items in the model. In general, loadings between 0·3 and 0·9 indicate an item is meaningfully related to the other items without overwhelming others in the model.^11,19^ Item thresholds characterize the location along the factor at which the cognitive test item provides maximal information of underlying cognitive function.

Loadings and thresholds/intercepts from the CFA models in HRS-HCAP were saved for use in serially estimated CFAs in subsequent HCAPs (**Supplemental Figure 1**). After estimating a CFA model for the HRS-HCAP study, we next estimated a CFA model in ELSA-HCAP, in which item parameters for cognitive test items in common with the HRS-HCAP were constrained to those observed in the HRS-HCAP, and the mean and variance of the underlying trait were freely estimated. The same process was repeated for all other HCAPs. Parameters for cognitive test items from a given HCAP study that were not yet in the item bank were freely estimated, then saved in the item bank for use when the next HCAP study was added to the item bank. In a final factor score-estimating CFA model for each cognitive domain, we estimated a CFA in the pooled sample of all HCAP studies, in which all parameters were fixed to previous values. We evaluated marginal reliability of the measurement models of each domain, calculated from the standard error of the measurement, in each HCAP.

#### Differential item functioning

The validity of the cross-national harmonization of cognitive function depends on the availability of common, equivalent cognitive test items across studies. While our expert panel identified equivalent linking items, it is possible to miss test differences that may have not been documented, that are due to unforeseen cultural differences, or for which there was insufficient documentation available. Thus, we statistically tested for differential item functioning (DIF) among candidate equivalent linking items between the HRS-HCAP and each study, by cognitive domain. We used multiple indicator, multiple cause (MIMIC) models to evaluate DIF by HCAP study membership.^15,24^ Briefly, we first tested DIF amongst cognitive test items rated as confident linking items. Next, we tested for DIF among cognitive items rated as tentative linking items, treating as anchors the confident items that showed no DIF in the prior analysis. The magnitude of DIF attributable to a given cognitive test item is represented by an odds ratio (OR) for an item on an indicator for study membership; we considered non-negligible DIF as an OR outside the range of 0·66 to 1·5.^25^ Large impact of DIF on participant’s domain-level scores, called salient DIF, was evaluated by taking the difference between DIF-adjusted and non-DIF-adjusted scores, via enabling items that showed DIF to have different measurement model parameters across studies, and counting how many participant scores would differ by more than 0·3 SD units.^26^

#### Validation

To evaluate construct validity, we evaluated the patterns of factor scores by age, gender, and educational attainment by regressing general cognitive function on each of these characteristics, adjusting for the other characteristics. We hypothesized better cognitive function on average at younger ages and with more educational attainment.^21,22^ With respect to gender, we hypothesized women are more disadvantaged in LMIC settings compared to men, given known gender-based societal inequalities in these settings that apply to determinants of later-life cognitive health such as educational opportunities.^23^

Descriptive analyses were conducted using Stata (Version 17, Stata Corp, College Station, TX). Factor analysis was conducted using Mplus.^27^

## Results

### Descriptive analyses

There were N=21,141 participants across the six HCAP studies (**Table 1**).

**Table 1.** Sample characteristics of included HCAP studies (N=21,141)

| Characteristics | Overall sample | HRS-HCAP | ELSA-HCAP | LASI-DAD | Mex-Cog | CHARLS - HCAP | HAALSI-HCAP |
| --- | --- | --- | --- | --- | --- | --- | --- |
| Sample size, n | 21141 | 3347 | 1273 | 4096 | 2042 | 9755 | 628 |
| Age, years, mean (SD) | 72.7 (8.9) | 76.6 (7.5) | 75.8 (7.0) | 69.7 (7.6) | 68.1 (9.0) | 68.5 (6.5) | 69.3 (11.5) |
| Female gender, n (%) | 16404 (55.7) | 2020 (60.4) | 700 (55.0) | 2207 (53.9) | 1203 (58.9) | 4960 (50.8) | 387 (61.6) |
| Education, n (%) |  |  |  |  |  |  |  |
| No or Early Childhood Education | 8862 (42.0) | 22 (0.7) | 3 (0.2) | 2558 (62.5) | 1023 (50.5) | 4909 (50.3) | 347 (55.3) |
| Primary education (US grades 1-6) | 3674 (17.4) | 131 (3.9) | 0 (0.0) | 527 (12.9) | 452 (22.3) | 2355 (24.1) | 209 (33.3) |
| Lower secondary (US grades 7-9) | 3160 (15.0) | 454 (13.6) | 486 (39.3) | 314 (7.7) | 317 (15.7) | 1562 (16.0) | 27 (4.3) |
| Upper secondary (US grades 10-12) | 3465 (16.4) | 1773 (53.0) | 303 (24.5) | 505 (12.3) | 60 (3.0) | 792 (8.1) | 32 (5.1) |
| Any college | 1924 (9.1) | 965 (28.8) | 446 (36.0) | 192 (4.7) | 172 (8.5) | 137 (1.4) | 12 (1.9) |

**Figure 1** (and **Supplemental Table 2**) displays the cognitive test items, stratified by cognitive domain to which tests were assigned. Of the 78 cognitive test items administered, 12 were judged by experts to be comparably administered in every HCAP. Overall, 15 distinct test items were assigned to the orientation domain, 14 distinct test items were assigned to the memory domain, 26 distinct cognitive test items were assigned to the executive functioning domain, and 23 distinct cognitive test items were assigned to the language domain. For a given test item in each column, the presence of factor loadings from the item banking approach reflect decisions about the comparability of items made during prestatistical harmonization. For example, for orientation, we determined that asking for one’s municipality in HAALSI-HCAP was comparable to asking for one’s district in LASI-DAD. Notably, our prestatistical team decided *a priori* that the CERAD word recall test was administered differently in the HRS-HCAP and ELSA-HCAP as compared to LASI-DAD, Mex-Cog, and HAALSI-HCAP because participants in the former two countries were presented with the words both verbally and visually, but in the latter three countries, participants were presented with the words only verbally. Moreover, while all studies presented the words verbally, there was variation in the order in which words were presented (i.e., alternating per trial vs fixed).

### HCAP-specific factor analyses

**Table 2** displays model fit statistics for measurement models of each of the five cognitive domains, by each of the seven study groups (six HCAP studies with LASI-DAD stratified by literacy). Of these 35 measurement models, 31% (11 models) were of perfect or good fit, 60% (21 models) were of adequate fit, and the remaining 9% (3 models) were of poor fit. Two of the three poorly fitting models were in the general cognitive function domain. Ultimately, we proceeded with these factor structures because most model fits were good or adequate.^28^

**Table 2.**
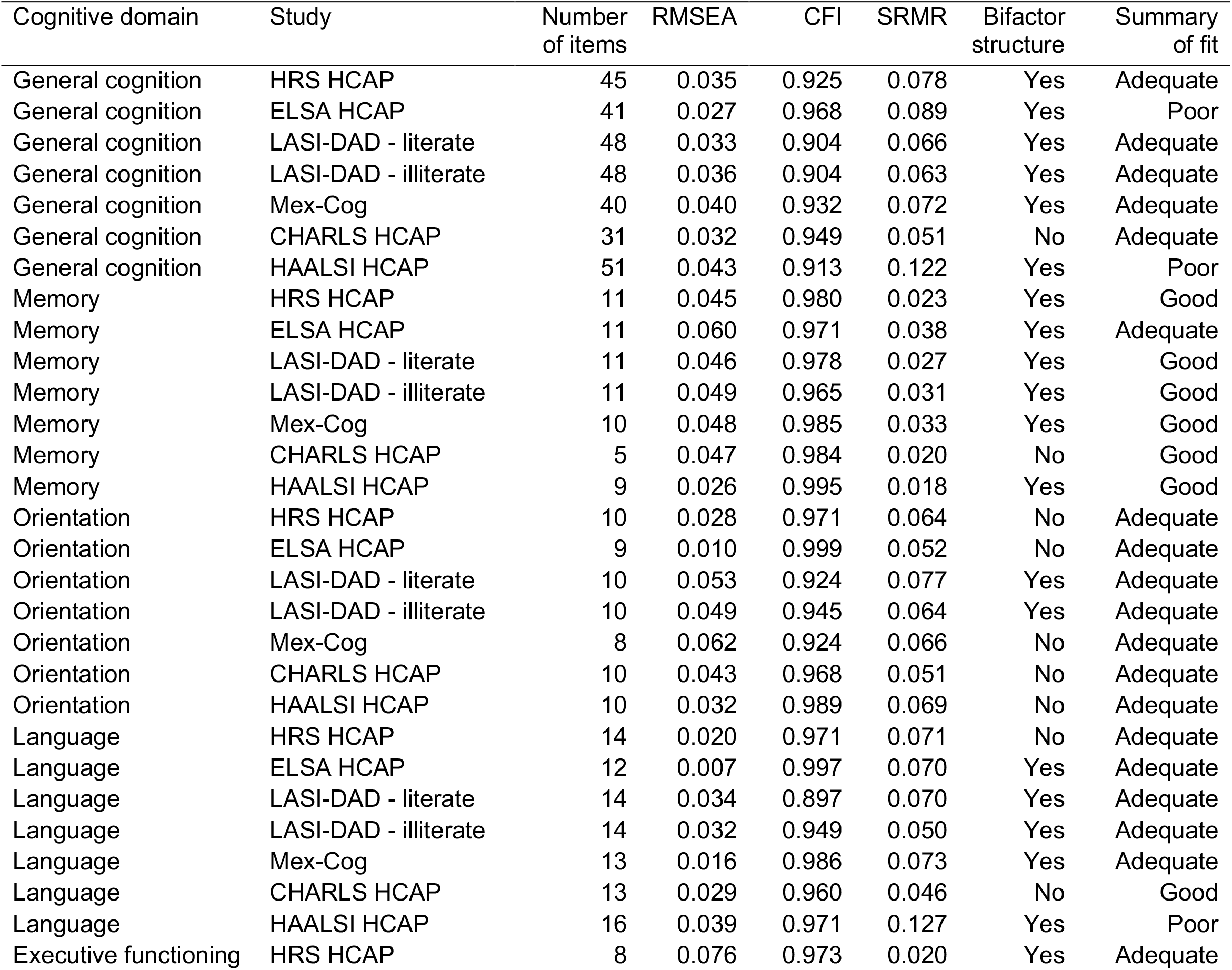
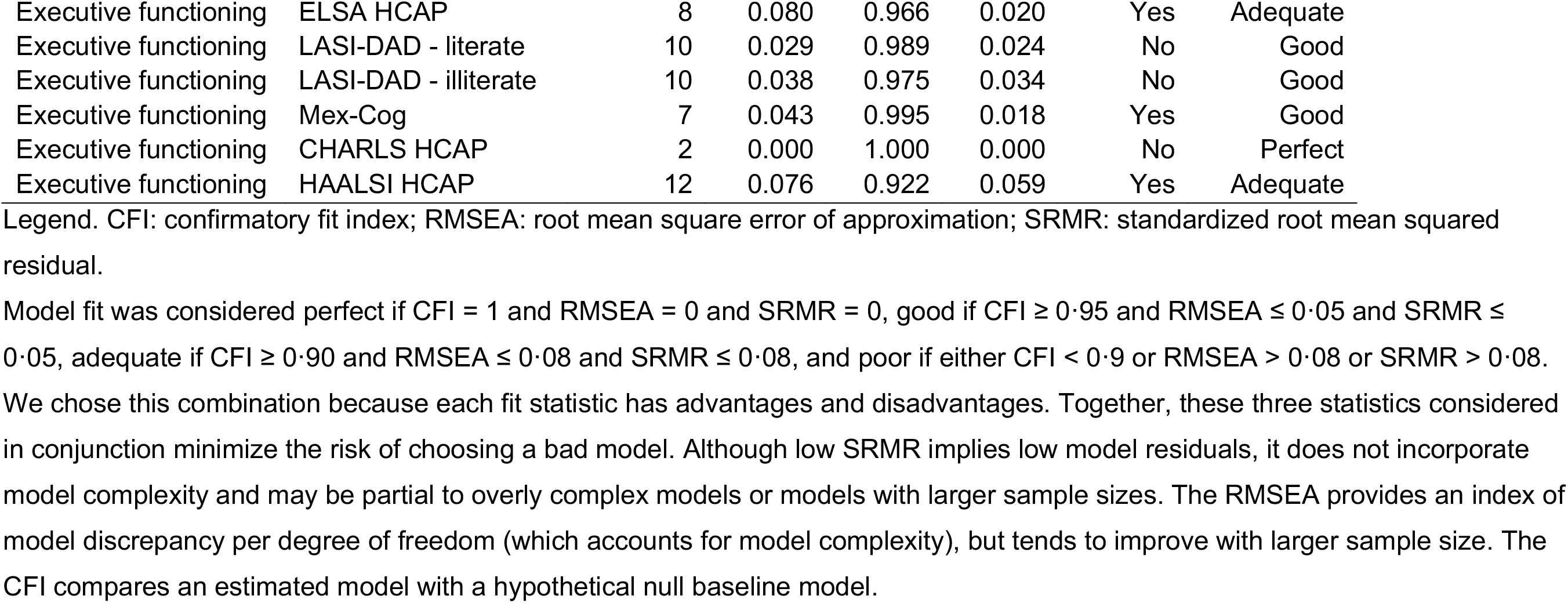
Model fit statistics of CFAs for each cognitive domain in each study: Results from HCAP studies (N=21,141)

### Statistical harmonization via item banking

The factor scores for general cognitive function, memory, language, and executive function were approximately normally distributed in each study (**Figure 2**). In contrast, the orientation factor showed a strong ceiling effect in each study **(Figure 2**). These ceiling effects are explained by low reliabilities (internal consistency based on the standard error of the measurement model) of the factor scores for the orientation domain. The orientation factor provided low precision above scores of 0 as more than half of participants in HRS-HCAP and a plurality of those in other studies were at the ceiling because they answered all orientation items correctly (**Figure 3**). In comparison, across a broad range of values, the reliabilities of the general cognitive function and memory factors are uniformly high (above r=0·9) for each HCAP between scores of -4 and 2, which encompasses over 90% of respondent scores for those domains. The language factor exhibited higher reliability at lower levels of language ability compared to higher levels, reflecting that almost all the language items, with the exception of animal fluency, tended to be easier questions about naming. For executive function, reliability was high for all studies except CHARLS-HCAP, which had just two test items measuring executive function.

**Figure 2.**
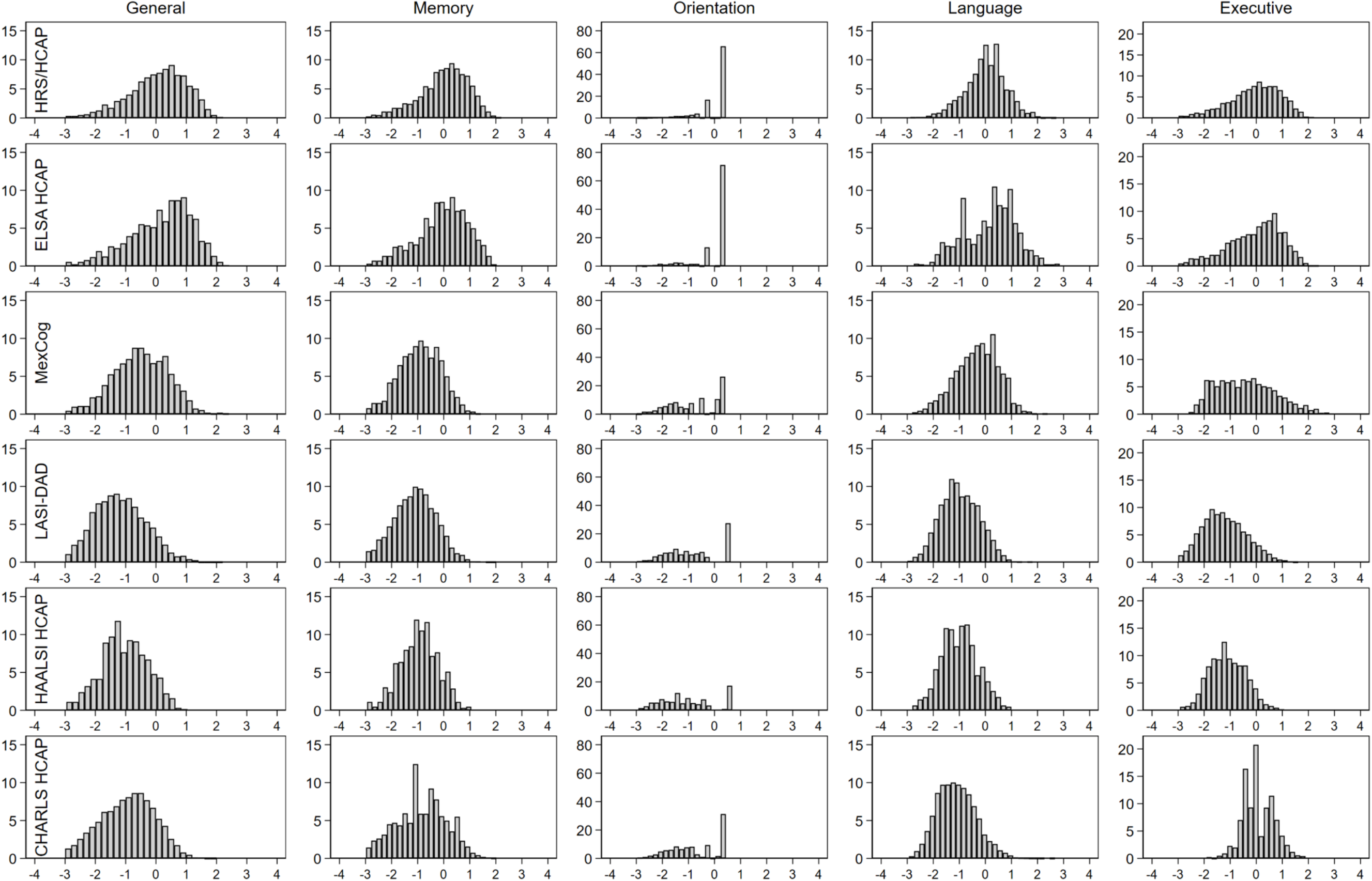
Distributions of harmonized general and domain-specific cognitive factor scores: Results from HCAP studies (N=21,141)

**Figure 3.**
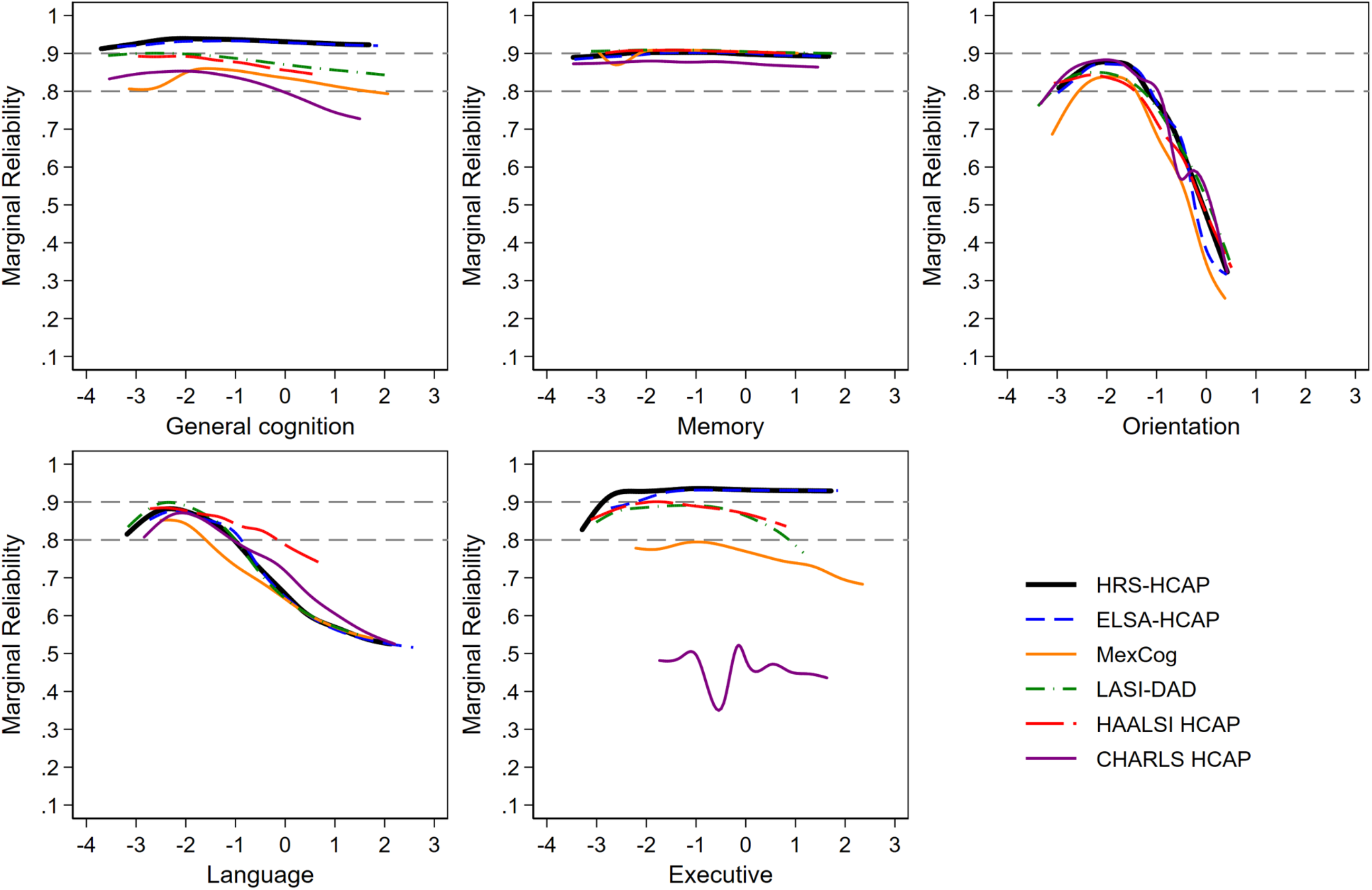
Plots of marginal reliability by study for overall and domain-specific cognitive performance: Results from HCAP studies (N=21,141)

### Differential item functioning

Evidence of DIF by study was present across cognitive test items (**Supplemental Table 3**). Of 78 unique cognitive test items, 23 showed DIF between HRS-HCAP and another study. Of these, 12 assessed language. With respect to the impact of DIF, 16·3% (N=290) of LASI-DAD orientation scores, 51·9% (N=326) of HAALSI-HCAP orientation scores, and 68·4% (N=6,668) of CHARLS-HCAP language factor score estimates demonstrated salient DIF (i.e., estimates differed by 0.3 units or more before vs. after accounting for DIF). Subsequent analyses, removing each item as a linking item one at a time, revealed that orientation to year was entirely responsible for all salient or impactful DIF in orientation for both LASI-DAD and HAALSI-HCAP (performance on this item was much lower in these studies, controlling for underlying orientation ability). Most of the salient DIF in CHARLS HCAP’s language domain could be attributed to differences in two items: naming a described cactus and reading and following a command. After removing these items as linking items between CHARLS HCAP and other studies (**Table 3**), 16·8% of participant scores were affected by adjustment for DIF in the language domain for CHARLS-HCAP. Otherwise, less than 6% of scores for any domain in any study was considerably impacted by DIF adjustment. **Figure 1** reflects decisions from DIF analyses to relax assumptions of item equivalence for orientation to year between LASI-DAD and HAALSI-HCAP with other studies, and for naming a described cactus and reading and following a command between CHARLS-HCAP and other studies.

**Table 3.**
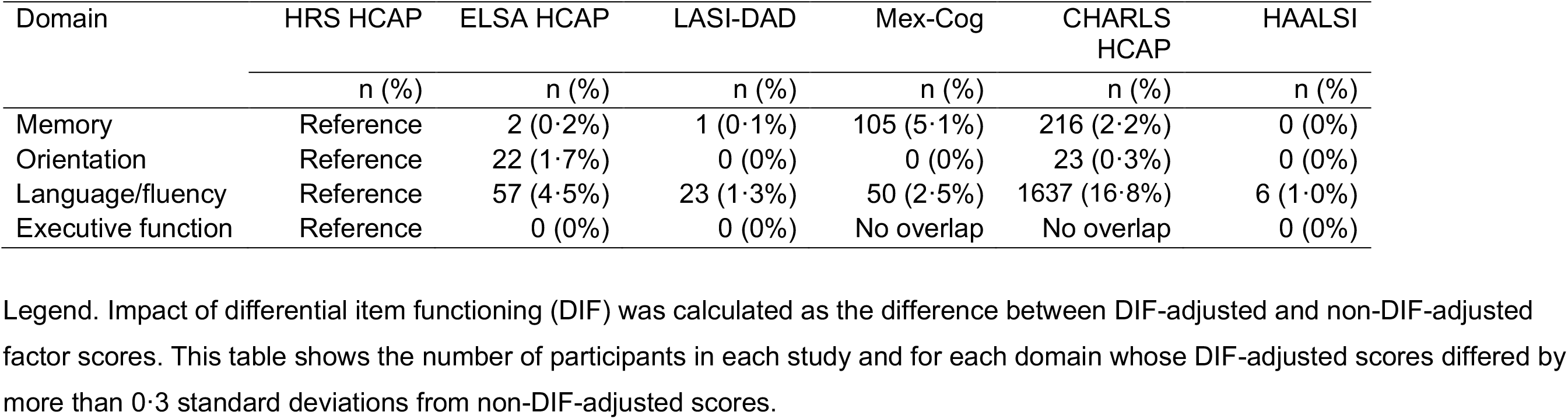
Number of participants in each study and for each domain whose scores show salient DIF: Results from HCAP studies (N=21,141)

| Domain | HRS HCAP | ELSA HCAP | LASI-DAD | Mex-Cog | CHARLS HCAP | HAALSI |
| --- | --- | --- | --- | --- | --- | --- |
|  | n (%) | n (%) | n (%) | n (%) | n (%) | n (%) |
| Memory | Reference | 2 (0.2%) | 1 (0.1%) | 105 (5.1%) | 216 (2.2%) | 0 (0%) |
| Orientation | Reference | 22 (1.7%) | 0 (0%) | 0 (0%) | 23 (0.3%) | 0 (0%) |
| Language/fluency | Reference | 57 (4.5%) | 23 (1.3%) | 50 (2.5%) | 1637 (16.8%) | 6 (1.0%) |
| Executive function | Reference | 0 (0%) | 0 (0%) | No overlap | No overlap | 0 (0%) |

Legend. Impact of differential item functioning (DIF) was calculated as the difference between DIF-adjusted and non-DIF-adjusted factor scores. This table shows the number of participants in each study and for each domain whose DIF-adjusted scores differed by more than 0.3 standard deviations from non-DIF-adjusted scores.

### Validation

Patterns of cognitive function aligned with hypothesized expectations: general cognitive function scores were, on average, lower at older ages and higher with greater education (**Table 4**). Women had higher average general cognitive function scores than men in the HRS-HCAP and ELSA-HCAP, but lower average scores than men in LASI-DAD, Mex-Cog, CHARLS-HCAP, and HAALSI-HCAP.

**Table 4.** Validation of the general cognitive function factor: Results from HCAP studies (N=21,141)

| Covariate | Overall sample | HRS-HCAP | ELSA-HCAP | LASI-DAD | Mex-Cog | CHARLS-HCAP | HAALSI-HCAP |
| --- | --- | --- | --- | --- | --- | --- | --- |
|  | Beta coefficient (SE) | Beta coefficient (SE) | Beta coefficient (SE) | Beta coefficient (SE) | Beta coefficient (SE) | Beta coefficient (SE) | Beta coefficient (SE) |
| Female gender | 0.017 (0.011) | 0.19 (0.03) | 0.06 (0.05) | -0.06 (0.03) | -0.10 (0.03) | -0.07 (0.02) | -0.22 (0.04) |
| Age group |  |  |  |  |  |  |  |
| 50-59 years | .40 (.043) | N/A | N/A | N/A | 0.28 (0.05) | N/A | 0.46 (0.07) |
| 60-69 years | REF | REF | REF | REF | REF | REF | REF |
| 70-79 years | .14 (.014) | -0.30 (0.04) | -0.57 (0.07) | -0.31 (0.03) | -0.54 (0.05) | -0.25 (0.02) | -0.38 (0.07) |
| 80-89 years | -.057 (.017) | -0.89 (0.04) | -1.23 (0.07) | -0.66 (0.04) | -1.17 (0.06) | -0.74 (0.04) | -0.72 (0.07) |
| 90+ years | -.34 (.032) | -1.45 (0.07) | -1.80 (0.14) | -1.18 (0.09) | -1.61 (0.14) | -0.99 (0.17) | -0.95 (0.14) |
| Education |  |  |  |  |  |  |  |
| No or Early Childhood education | -1.16 (0.017) | -0.55 (0.20) | -1.01 (0.58) | -0.96 (0.04) | -1.26 (0.05) | -1.13 (0.02) | -0.96 (0.12) |
| Primary education (US grades 1-6) | -0.36 (0.019) | -0.50 (0.09) | N/A | -0.26 (0.05) | -0.42 (0.05) | -0.35 (0.03) | -0.32 (0.12) |
| Lower secondary (US grades 7-9) | REF | REF | REF | REF | REF | REF | REF |
| Upper secondary (US grades 10-12) | 0.40 (0.020) | 0.72 (0.05) | 0.52 (0.07) | 0.34 (0.05) | 0.17 (0.10) | 0.26 (0.03) | 0.47 (0.16) |
| Any college | 0.84 (0.023) | 1.18 (0.05) | 0.76 (0.07) | 0.55 (0.06) | 0.57 (0.07) | 0.50 (0.07) | 0.78 (0.21) |

Legend. Beta coefficients represent overall and study-specific differences in general cognitive functioning between a given exposure grouping and the reference category. For age, persons aged 60-69 comprised the reference group. For education, persons with a lower secondary education comprised the reference group.

## Discussion

We investigated the performance of common and unique cognitive test items administered to 21,141 older adults across six large harmonized studies of aging in the US, England, India, Mexico, China, and South Africa. We demonstrated these cognitive test items empirically reflect comparable domains of cognitive function among older adults living across these countries, they are reliable and valid measures of cognitive function, and useful for population-based research. Most importantly, we overcame differences in test administration due to language, literacy, and numeracy to statistically harmonize general and domain-specific cognitive function across these countries.

Over the past decade, a growing number of cross-national studies have examined risk factors of cognitive function decline and dementia, mostly using data from the HRS and its IPS.^29–33^ Risk factors examined in these studies have included socioeconomic characteristics, health behaviors, physical and mental health conditions, and telomere length.^21,34–55^ However, none of them conducted in-depth statistical harmonization of cognitive test items.

High-quality, harmonized scores for general and domain-specific cognitive function are crucial tools to promote valid cross-national comparisons of predictors and outcomes of cognitive aging in a rapidly aging world.^17^ A recent Lancet Commission report identified 12 risk factors that had strong evidence of a causal risk for dementia: low education, hearing impairment, traumatic brain injury, hypertension, diabetes, excessive alcohol use, obesity, smoking, depression, social isolation, physical inactivity, and air pollution.^56^ The harmonized cognitive function scores generated here can be used in pooled analyses to evaluate whether these risk factors have similar effects on cognitive function across global settings. Such knowledge could facilitate identification of contextual risk-modifying factors that could be intervened upon to reduce the risk of dementia in certain populations.^57^ Further, common cognitive phenotypes could be used to improve the quality of population attributable risk estimates. Finally, common cognitive phenotypes could be used in dementia algorithms that are applied cross-nationally to generate prevalence and incidence estimates that are truly comparable across national settings.^58^

Prestatistical harmonization accompanied by statistical testing for differential item functioning (DIF) were two essential steps to the harmonization goal of this study. DIF can be introduced by methodological differences in test administration or scoring across studies, in addition to population-level differences that may alter responses to equivalent test items (e.g., differences in literacy, numeracy, and language). We evaluated the comparability of cognitive test items using a multidisciplinary team, which was a crucial component of this harmonization work. However, an expert’s ability to identify measurement differences in cognitive test items across languages and cultures in items depends on the quality of available study documentation and level of expertise regarding the population under study. We are confident in our pre-statistical harmonization given the available documentation and our team’s level of expertise, but adequate documentation is crucial. Statistical DIF testing identified only four of 20 domain-by-country categories in which the DIF made a difference for more than 10% of the sample, however in these cases the DIF proved critical to the estimation of scores.

Strengths of this study include nationally representative sampling (in the case of HAALSI, regionally representative sampling) and comprehensive cognitive phenotyping with a common protocol. All data are publicly available (see **Supplemental Table 1**). Our harmonization approach based on item banking is readily scalable: as data from more HCAPs are released or become available, and as longitudinal data from existing HCAPs become available, they can be readily added to our item bank to be harmonized alongside the data shown here. Alongside these strengths, there are notable limitations. The quality of the linking between studies is best when there are more cognitive test items with richer distributions. This poses challenges when domains largely include relatively easy dichotomous items (e.g. language and orientation). A further limitation is that while we identified DIF, it was outside the scope of this study to characterize possible reasons for DIF across HCAP batteries in each item. This is a worthwhile aim for future research, especially as test batteries are adapted to additional countries and contexts.

In conclusion, the HCAP suite of cognitive test items administered in the US, England, India, Mexico, China, and South Africa reflects a common structure of general- and domain-specific cognitive function across these diverse countries. Despite common protocols, there were necessary item adaptations to account for language, literacy, numeracy, and cultural differences across participating countries. Statistical harmonization involving an item-banking approach with identification of common and unique items allowed for the construction of reliable and valid factor scores that account for these differences. Future cross-national comparisons of risk factors for cognitive aging outcomes, estimates of dementia prevalence and incidence, and estimates of population attributable fractions of risk factors should consider using harmonized factor scores to improve the quality of their analyses.

## Data Availability

The following are the links to which we used to access the data. Most of the studies we used were available on the "Gateway to Global Aging" (g2aging.org) website, except for the HAALSI dataset. All datasets used were publicly available data. All necessary data use agreements were signed and submitted by study team members.
HRS HCAP: https://hrs.isr.umich.edu/data-products/hcap
ELSA HCAP: https://beta.ukdataservice.ac.uk/datacatalogue/series/series?id=200011#!/access-data
Mex-Cog: https://www.mhasweb.org/DataProducts/HarmonizedData.aspx
CHARLS HCAP: http://charls.pku.edu.cn/en/Data/Harmonized_CHARLS.htm
HAALSI HCAP: https://haalsi.org/data
LASI-DAD: https://g2aging.org/?section=lasi-downloads#download-listing-lasi-wave-1

https://beta.ukdataservice.ac.uk/datacatalogue/series/series?id=200011#!/access-data

https://www.mhasweb.org/DataProducts/HarmonizedData.aspx

http://charls.pku.edu.cn/en/Data/Harmonized_CHARLS.htm

https://g2aging.org/?section=lasi-downloads#download-listing-lasi-wave-1

https://haalsi.org/data

https://g2aging.org/downloads

