## supplemental tables and figures for "Harmonization of Later-Life Cognitive Function Across National Contexts: Results from the Harmonized Cognitive Assessment Protocols (HCAPs)"

Supplemental Table 1. Information about HCAP studies

| | United States | England | India | Mexico | China | South Africa |
|---|---|---|---|---|---|---|
| Parent cohort study | Health and Retirement study (HRS) | English Longitudinal Study on Ageing (ELSA) | Longitudinal Aging Study in India (LASI) | Mexican Health and Aging Study (MHAS) | China Health and Retirement Longitudinal Study (CHARLS) | Health and Aging in Africa: A Longitudinal Study of an INDEPTH Community in South Africa (HAALSI) |
| HCAP sub-study | Harmonized Cognitive Assessment Protocol Project of Health and Retirement study (HRS-HCAP) | Harmonised Cognitive Assessment Protocol Sub-study of the English Longitudinal Study of Ageing (ELSA-HCAP) | Harmonised Diagnostic Assessment of Dementia for the Longitudinal Aging Study in India (LASI-DAD) | Mexican Cognitive Aging Ancillary Study (Mex-Cog) | Harmonized Cognitive Assessment Protocol for the China Health and Retirement Longitudinal Study (CHARLS-HCAP) | Cognition and dementia in the Health and Aging in Africa Longitudinal Study of an INDEPTH community in South Africa (HAALSI-HCAP) |
| Year in HCAPs implemented | 2016 | 2018 | 2017-2019 | 2016 | 2018 | 2019 |
| Eligibility criteria for HCAPs | 1. Aged 65 years and over at the time of HRS-HCAP survey<br>2. Completed core interview | 1. Aged 65 years and over at the time of ELSA-HCAP survey<br>2. Completed an ELSA interview in wave 7 (2014-15) or wave 8 (2016-17) | 1. Aged 60 years and over at the time of LASI-DAD survey | 1. Aged 55 and over in the MHAS 2015<br>2. Completed a direct or proxy interview for health reasons in the MHAS 2015 | 1. Aged 60 and over at the time of CHARLS-HCAP | 1. Aged 50 and over |
| Number of target cases to conduct HCAPs | 4,425 | 1,778 | 3,891 | 3,250 | Not reported | Not reported |

| | | | | | | |
|---|---|---|---|---|---|---|
| Number of completed HCAPs | 3,496 | 1,273 | 4,096 | 2,042 | 9,755 | 628 |

Supplemental Table 2. Cognitive test items and their overlap across each study: Results from HCAP studies (N=21,141)

| Variable | HRS-HCAP | ELSA-HCAP | LASI-DAD (literate) | LASI-DAD (illiterate) | Mex-Cog | CHARLS-HCAP | HAALSI-HCAP |
|---|---|---|---|---|---|---|---|
| **Orientation** | | | | | | | |
| Day of month | 0·68 (0·43) | 0·68 (0·43) | 0·68 (0·43) | 0·68 (0·43) | 0·68 (0·43) | 0·68 (0·43) | 0·68 (0·43) |
| Month | 0·88 (0·73) | 0·88 (0·73) | 0·88 (0·73) | 0·88 (0·73) | 0·88 (0·73) | 0·88 (0·73) | 0·88 (0·73) |
| Year | 0·90 (0·76) | 0·90 (0·76) | | | 0·90 (0·76) | 0·90 (0·76) | |
| Year (DIF adjusted) | | | 0·70 (0·82) | 0·70 (0·82) | | | 0·70 (0·82) |
| Day of the week | 0·76 (0·63) | 0·76 (0·63) | 0·76 (0·63) | 0·76 (0·63) | 0·76 (0·63) | 0·76 (0·63) | 0·76 (0·63) |
| What time is it | | | | | 0·17 (0·32) | | 0·17 (0·32) |
| Where are we | | | | | 0·13 (0·35) | | |
| What country are we in | | 0·62 (0·72) | | | 0·62 (0·72) | | 0·62 (0·72) |
| What state are we in | 0·64 (0·61) | | 0·64 (0·61) | 0·64 (0·61) | 0·64 (0·61) | 0·64 (0·61) | 0·64 (0·61) |
| What county are we in | 0·68 (0·55) | 0·68 (0·55) | | | | 0·68 (0·55) | |
| What city are we in | 0·83 (0·72) | 0·83 (0·72) | 0·83 (0·72) | 0·83 (0·72) | | 0·83 (0·72) | 0·83 (0·72) |
| Season of year | 0·54 (0·42) | 0·54 (0·42) | 0·54 (0·42) | 0·54 (0·42) | | 0·54 (0·42) | 0·54 (0·42) |
| Floor of building | 0·66 (0·56) | | 0·66 (0·56) | 0·66 (0·56) | | 0·66 (0·56) | 0·66 (0·56) |
| Address (street name and/or building number) | 0·76 (0·62) | 0·76 (0·62) | 0·76 (0·62) | 0·76 (0·62) | | 0·76 (0·62) | |
| Name of hospital or district/municipality | | | 0·63 (0·69) | 0·63 (0·69) | | | 0·63 (0·69) |
| **Memory** | | | | | | | |
| CERAD immediate sum of 3 trials | 0·87 (0·82) | 0·87 (0·82) | | | | 0·87 (0·82) | |
| CERAD immediate sum of 3 trials | | | 0·89 (0·69) | 0·89 (0·69) | 0·89 (0·69) | | 0·89 (0·69) |
| CERAD word list delay | 0·88 (0·85) | 0·88 (0·85) | | | | 0·88 (0·85) | |
| CERAD word list delay | | | 0·88 (0·65) | 0·88 (0·65) | 0·88 (0·65) | | 0·88 (0·65) |
| CERAD recognition | 0·75 (0·58) | 0·75 (0·58) | | | | 0·75 (0·58) | |
| CERAD recognition | | | 0·79 (0·65) | 0·79 (0·65) | 0·79 (0·65) | | 0·79 (0·65) |
| Three word immediate registration | 0·51 (0·49) | 0·51 (0·49) | 0·51 (0·49) | 0·51 (0·49) | 0·51 (0·49) | 0·51 (0·49) | 0·51 (0·49) |
| Three word delayed recall | 0·76 (0·65) | 0·76 (0·65) | 0·76 (0·65) | 0·76 (0·65) | 0·76 (0·65) | 0·76 (0·65) | 0·76 (0·65) |
| Logical Memory immediate | 0·71 (0·65) | 0·71 (0·65) | 0·71 (0·65) | 0·71 (0·65) | 0·71 (0·65) | | 0·71 (0·65) |
| Logical Memory delay | 0·74 (0·67) | 0·74 (0·67) | 0·74 (0·67) | 0·74 (0·67) | 0·74 (0·67) | | 0·74 (0·67) |
| Logical memory recognition | 0·62 (0·52) | 0·62 (0·52) | 0·62 (0·52) | 0·62 (0·52) | | | 0·62 (0·52) |
| Brave man immediate (East Boston Memory Test) | 0·42 (0·39) | 0·42 (0·39) | 0·42 (0·39) | 0·42 (0·39) | 0·42 (0·39) | | |
| Brave man delay (East Boston Memory Test) | 0·53 (0·43) | 0·53 (0·43) | 0·53 (0·43) | 0·53 (0·43) | 0·53 (0·43) | | |
| CERAD constructional praxis delay | 0·70 (0·67) | 0·70 (0·67) | 0·70 (0·67) | 0·70 (0·67) | 0·70 (0·67) | | 0·70 (0·67) |
| **Executive functioning** | | | | | | | |

| | | | | | | | |
|---|---|---|---|---|---|---|---|
| Problem solving | | | 0·75 (0·71) | 0·75 (0·71) | | | |
| Ravens progressive matrices | 0·74 (0·68) | 0·74 (0·68) | 0·74 (0·68) | 0·74 (0·68) | | | 0·74 (0·68) |
| HRS Number series | 0·64 (0·57) | 0·64 (0·57) | | | | | |
| Number series | | | | | | 0·56 (0·44) | |
| Trails A time (letters and numbers) | 0·79 (0·76) | 0·79 (0·76) | | | | | |
| Trails B time (letters and numbers) | 0·76 (0·67) | 0·76 (0·67) | | | | | |
| Similarities | | | 0·58 (0·57) | 0·58 (0·57) | 0·58 (0·57) | | 0·58 (0·57) |
| Token Test | | | 0·77 (0·75) | 0·77 (0·75) | | | 0·77 (0·75) |
| Digit Span Forward (single item) | | | 0·68 (0·60) | 0·68 (0·60) | | | |
| Digit Span Backward (single item) | | | 0·73 (0·64) | 0·73 (0·64) | | | |
| Digit Span Forward (multiple items) | | | | | | | 0·48 (0·38) |
| Digit Span Backward (multiple items) | | | | | | | 0·43 (0·28) |
| Go-No-Go | | | 0·71 (0·67) | 0·71 (0·67) | 0·71 (0·67) | | 0·71 (0·67) |
| Motor Programming | | | | | | | 0·76 (0·74) |
| MMSE Spelling backwards | 0·62 (0·65) | | | | | | |
| Backward counting, 100-0 | 0·69 (0·63) | 0·69 (0·63) | | | | | |
| Backward counting, 20-0 | | | | | 0·54 (0·79) | | |
| Symbol Digit Modalities Test * | 0·88 (0·77) | 0·88 (0·77) | | | | | |
| Symbols and Digits test ** | | | | | 0·58 (0·77) | | |
| Symbol Cancellation Test | | | 0·67 (0·68) | 0·67 (0·68) | 0·67 (0·68) | | 0·67 (0·68) |
| Letter cancellation | 0·59 (0·57) | 0·59 (0·57) | | | | | |
| Serial 3s | | | | | 0·36 (0·54) | | |
| Serial 7s | | 0·56 (0·53) | 0·56 (0·53) | 0·56 (0·53) | 0·56 (0·53) | 0·56 (0·53) | 0·56 (0·53) |
| Backward Day naming | | | 0·68 (0·68) | 0·68 (0·68) | | | 0·68 (0·68) |
| Forward day naming | | | | | | | 0·74 (0·68) |
| CDR calculation-cent | | | | | | | 0·62 (0·69) |
| **Language** | | | | | | | |
| Animal fluency | 0·74 (0·68) | 0·74 (0·68) | 0·74 (0·68) | 0·74 (0·68) | 0·74 (0·68) | 0·74 (0·68) | 0·74 (0·68) |
| Name a described cactus | 0·81 (0·70) | 0·81 (0·70) | | | | | |
| Name a described cactus (DIF adjusted) | | | | | | 0·79 (0·60) | |
| Name a described coconut | | | 0·56 (0·45) | 0·56 (0·45) | | | |
| What are scissors used for? | 0·71 (0·54) | 0·71 (0·54) | 0·71 (0·54) | 0·71 (0·54) | 0·71 (0·54) | 0·71 (0·54) | 0·71 (0·54) |
| Object naming (watch) | 0·69 (0·59) | | 0·69 (0·59) | 0·69 (0·59) | 0·69 (0·59) | 0·69 (0·59) | 0·69 (0·59) |
| Object naming (pencil) | 0·56 (0·57) | | 0·56 (0·57) | 0·56 (0·57) | 0·56 (0·57) | 0·56 (0·57) | 0·56 (0·57) |
| Name the elbow | 0·88 (0·68) | 0·88 (0·68) | 0·88 (0·68) | 0·88 (0·68) | 0·88 (0·68) | 0·88 (0·68) | 0·88 (0·68) |
| Write a sentence (or write one's name) | 0·62 (0·52) | 0·62 (0·52) | 0·62 (0·52) | | 0·62 (0·52) | 0·62 (0·52) | 0·62 (0·52) |

| | | | | | | | |
|---|---|---|---|---|---|---|---|
| Say a sentence | | | | 0·81 (0·67) | | | |
| Read and follow command (Close your eyes) | 0·61 (0·46) | 0·61 (0·46) | 0·61 (0·46) | | 0·61 (0·46) | | 0·61 (0·46) |
| Read and follow command (DIF adjusted) | | | | | | 0·73 (0·58) | |
| Follow example (close your eyes) | | | | 0·86 (0·71) | | | |
| Repetition of a phrase | 0·46 (0·37) | 0·46 (0·37) | 0·46 (0·37) | 0·46 (0·37) | 0·46 (0·37) | 0·46 (0·37) | 0·46 (0·37) |
| What does one do with a hammer | 0·43 (0·24) | 0·43 (0·24) | 0·43 (0·24) | 0·43 (0·24) | 0·43 (0·24) | 0·43 (0·24) | 0·43 (0·24) |
| Define Bridge | | | | | 0·62 (0·54) | | |
| Point to 2 things in the vicinity | 0·85 (0·65) | 0·85 (0·65) | 0·85 (0·65) | 0·85 (0·65) | 0·85 (0·65) | 0·85 (0·65) | 0·85 (0·65) |
| Where is the local market? | 0·58 (0·48) | 0·58 (0·48) | 0·58 (0·48) | 0·58 (0·48) | 0·58 (0·48) | 0·58 (0·48) | 0·58 (0·48) |
| Follow 3-stage instruction | 0·39 (0·30) | 0·39 (0·30) | 0·39 (0·30) | 0·39 (0·30) | 0·39 (0·30) | | 0·39 (0·30) |
| Name president or Prime Minister | 0·85 (0·82) | 0·85 (0·82) | 0·85 (0·82) | 0·85 (0·82) | | 0·85 (0·82) | 0·85 (0·82) |
| Name deputy president | | | | | | | 0·87 (0·75) |
| Phonemic Fluency | | | | | | | 0·59 (0·63) |
| Boston Naming Test, uncued | | | | | | | 0·68 (0·62) |

Legend. This table shows factor loadings for domain-specific factor analyses, and in parentheses the factor loadings for the model for general cognitive function. Loadings are standardized to have a range from -1 to 1, thus can be interpretable as correlations between items and the underlying factor. The presence of factor loadings for a given test item in each column reflects decisions about the comparability of items made at the prestatistical harmonization as well as after testing for differential item functioning. Refer to the Methods and Results for details.

* (110 items, fill in number for a given symbol)

** (56 items, fill in symbol for a given number)

Supplemental Table 3. Results of differential item functioning among confident and tentative linking items: Results from HCAP (N=21,141)

| Study | Cognitive domain | Stage of DIF testing | Cognitive test item | Association with cohort (odds ratio) [a] | 95% CI lower bound | 95% CI [b] upper bound | Interpretation |
|---|---|---|---|---|---|---|---|
| HAALSI HCAP | Language | | | | | | |
| | | DIF among confident linking items | | | | | |
| | | | Name the elbow | 1.65 | 1.31 | 2.08 | DIF |
| | | | Animal fluency | N/A | | | No DIF |
| | | | What are scissors used for? | N/A | | | No DIF |
| | | | Point to 2 things in the vicinity | N/A | | | No DIF |
| | | | Name president or Prime Minister | N/A | | | No DIF |
| | | DIF among tentative linking items, treating confident items as anchors | | | | | |
| | | | What does one do with a hammer | 2.64 | 2.14 | 3.27 | DIF |
| | | | Where is the local market? | 2.06 | 1.81 | 2.34 | DIF |
| ELSA HCAP | Language | | | | | | |
| | | DIF among confident linking items | | | | | |
| | | | Name a described cactus | 0.61 | 0.55 | 0.67 | DIF |
| | | | What does one do with a hammer | 2.29 | 1.86 | 2.82 | DIF |
| | | | Follow 3-stage instruction | 1.83 | 1.66 | 2.03 | DIF |
| | | | Name president or Prime Minister | 0.43 | 0.39 | 0.47 | DIF |
| | | | Animal fluency | N/A | | | No DIF |
| | | | Point to 2 things in the vicinity | N/A | | | No DIF |
| | | | What are scissors used for? | N/A | | | No DIF |
| | | | Name the elbow | N/A | | | No DIF |
| | | | Read and follow command (Close your eyes) | N/A | | | No DIF |
| | | | Repetition of a phrase | N/A | | | No DIF |
| | | DIF among tentative linking items, treating confident items as anchors | | | | | |
| | | | Where is the local market? | 2.31 | 2.03 | 2.62 | DIF |

| | | | | | | |
|---|---|---|---|---|---|---|
| | | Write a sentence (or write one's name) | N/A | | | No DIF |
| LASI-DAD | Language | | | | | |
| | | DIF among confident linking items | | | | |
| | | What are scissors used for? | N/A | | | No DIF |
| | | Object naming (pencil) | N/A | | | No DIF |
| | | Write a sentence (or write one's name) | 1.40 | 1.24 | 1.58 | Negligible |
| | | Read and follow command (Close your eyes) | 0.14 | 0.13 | 0.16 | DIF |
| | | Follow 3-stage instruction | 1.65 | 1.51 | 1.80 | DIF |
| | | Animal fluency | N/A | | | No DIF |
| | | Object naming (watch) | N/A | | | No DIF |
| | | Name the elbow | 1.13 | 0.96 | 1.32 | Negligible |
| | | Point to 2 things in the vicinity | N/A | | | No DIF |
| | | Name president or Prime Minister | N/A | | | No DIF |
| | | DIF among tentative linking items, treating confident items as anchors | | | | |
| | | Repetition of a phrase | 4.85 | 4.32 | 5.44 | DIF |
| | | What does one do with a hammer | N/A | | | No DIF |
| | | Where is the local market? | 3.79 | 3.36 | 4.27 | DIF |
| Mex-Cog | Language | | | | | |
| | | DIF among confident linking items | | | | |
| | | Animal fluency | N/A | | | No DIF |
| | | Object naming (pencil) | N/A | | | No DIF |
| | | Name the elbow | 1.03 | 0.87 | 1.21 | Negligible |
| | | Read and follow command (Close your eyes) | N/A | | | No DIF |
| | | Point to 2 things in the vicinity | N/A | | | No DIF |
| | | Follow 3-stage instruction | N/A | | | No DIF |
| | | DIF among tentative linking items, treating confident items as anchors | | | | |
| | | What are scissors used for? | 1.71 | 1.42 | 2.06 | DIF |
| | | Repetition of a phrase | 3.21 | 2.93 | 3.51 | DIF |

| | | | | | | | |
|---|---|---|---|---|---|---|---|
| | | | What does one do with a hammer | 2.08 | 1.83 | 2.37 | DIF |
| | | | Where is the local market? | 0.48 | 0.45 | 0.52 | DIF |
| | | | Object naming (watch) | N/A | | | |
| | | | Write a sentence (or write one's name) | N/A | | | |
| CHARLS HCAP | Language | | | | | | |
| | | DIF among confident linking items | | | | | |
| | | | Name the elbow | 0.44 | 0.39 | 0.50 | DIF |
| | | | Write a sentence (or write one's name) | 0.53 | 0.48 | 0.58 | DIF |
| | | | What does one do with a hammer | 2.32 | 2.12 | 2.54 | DIF |
| | | | Point to 2 things in the vicinity | 0.43 | 0.37 | 0.49 | DIF |
| | | | Name president or Prime Minister | 1.40 | 1.29 | 1.52 | Negligible |
| | | DIF among tentative linking items, treating confident items as anchors | | | | | |
| | | | Object naming (watch) | 0.59 | 0.49 | 0.70 | DIF |
| | | | Object naming (pencil) | N/A | | | |
| | | | Where is the local market? | 1.47 | 1.37 | 1.57 | Negligible |
| HAALSI HCAP | Memory | | | | | | |
| | | DIF among confident linking items | | | | | |
| | | | None available | | | | |
| | | DIF among tentative linking items, with no anchors | | | | | |
| | | | Logical Memory immediate | N/A | | | |
| | | | Logical Memory delay | 3.49 | 3.01 | 3.97 | DIF |
| | | | CERAD constructional praxis delay | -1.76 | -2.10 | -1.43 | DIF |
| | | | Logical memory recognition | -0.17 | -0.46 | 0.12 | DIF |
| ELSA HCAP | Memory | | | | | | |
| | | DIF among confident linking items | | | | | |
| | | | Three word delayed recall | 0.51 | 0.48 | 0.54 | Negligible |
| | | | Three word immediate registration | 1.43 | 1.25 | 1.63 | Negligible |
| | | | CERAD immediate sum of 3 trials | N/A | | | No DIF |

| | | | | | | |
|---|---|---|---|---|---|---|
| | | CERAD word list delay | N/A | | | No DIF |
| | | CERAD recognition | N/A | | | No DIF |
| | DIF among tentative linking items, treating confident items as anchors | | | | | |
| | | Logical Memory immediate | -2.05 | -2.27 | -1.82 | DIF |
| | | Logical memory recognition | 0.45 | 0.30 | 0.60 | DIF |
| | | Brave man delay (East Boston Memory Test) | 1.68 | 1.58 | 1.78 | DIF |
| | | Brave man immediate (East Boston Memory Test) | 2.29 | 2.17 | 2.42 | DIF |
| | | Logical Memory delay | N/A | | | No DIF |
| LASI-DAD | Memory | | | | | |
| | DIF among confident linking items | | | | | |
| | | Logical Memory delay | -0.34 | -0.56 | -0.12 | DIF |
| | | Three word delayed recall | 0.86 | 0.81 | 0.92 | Negligible |
| | | Brave man delay (East Boston Memory Test) | 1.83 | 1.73 | 1.93 | DIF |
| | | Brave man immediate (East Boston Memory Test) | 1.58 | 1.49 | 1.66 | DIF |
| | | Three word immediate registration | N/A | | | No DIF |
| | | Logical Memory immediate | N/A | | | No DIF |
| | | Logical memory recognition | N/A | | | No DIF |
| | DIF among tentative linking items, treating confident items as anchors | | | | | |
| | | CERAD constructional praxis delay | -0.89 | -1.07 | -0.72 | DIF |
| Mex-Cog | Memory | | | | | |
| | DIF among confident linking items | | | | | |
| | | CERAD constructional praxis delay | N/A | | | No DIF |
| | DIF among tentative linking items, treating confident items as anchors | | | | | |
| | | Logical Memory immediate | -2.72 | -2.94 | -2.49 | DIF |
| | | Logical Memory delay | -1.73 | -1.98 | -1.47 | DIF |
| | | Three word delayed recall | 0.80 | 0.75 | 0.86 | Negligible |
| | | Brave man delay (East Boston Memory Test) | 1.69 | 1.60 | 1.80 | DIF |
| | | Brave man immediate (East Boston Memory Test) | N/A | | | No DIF |

| | | | | | | |
|---|---|---|---|---|---|---|
| | | Three word immediate registration | N/A | | | No DIF |
| CHARLS HCAP | Memory | | | | | |
| | DIF among confident linking items | | | | | |
| | | Three word delayed recall | N/A | | | No DIF |
| | | Three word immediate registration | N/A | | | No DIF |
| | DIF among tentative linking items, treating confident items as anchors | | | | | |
| | | CERAD immediate sum of 3 trials | -0.14 | -0.35 | 0.07 | Negligible |
| | | CERAD word list delay | 0.92 | 0.82 | 1.01 | Negligible |
| | | CERAD recognition | 0.87 | 0.73 | 1.01 | Negligible |
| HAALSI HCAP | Orientation | | | | | |
| | DIF among confident linking items | | | | | |
| | | What state are we in | 0.16 | 0.13 | 0.19 | DIF |
| | | Day of the week | N/A | | | No DIF |
| | | What city are we in | N/A | | | No DIF |
| | DIF among tentative linking items, treating confident items as anchors | | | | | |
| | | Floor of building | N/A | | | No DIF |
| | | Season of year | N/A | | | No DIF |
| ELSA HCAP | Orientation | | | | | |
| | DIF among confident linking items | | | | | |
| | | Day of month | N/A | | | No DIF |
| | | Month | N/A | | | No DIF |
| | | Year | N/A | | | No DIF |
| | | Day of the week | N/A | | | No DIF |
| | | What city are we in | N/A | | | No DIF |
| | DIF among tentative linking items, treating confident items as anchors | | | | | |
| | | Season of year | 1.66 | 1.48 | 1.86 | DIF |
| | | Address | 1.38 | 1.20 | 1.59 | Negligible |
| | | What county are we in | N/A | | | No DIF |

| | | | | | | | |
|---|---|---|---|---|---|---|---|
| LASI-DAD | Orientation | | | | | | |
| | | DIF among confident linking items | | | | | |
| | | | Day of month | 1.35 | 1.23 | 1.48 | Negligible |
| | | | Day of the week | 0.81 | 0.73 | 0.90 | Negligible |
| | | | What state are we in | 0.28 | 0.24 | 0.32 | DIF |
| | | | Season of year | 1.47 | 1.33 | 1.63 | Negligible |
| | | | Month | N/A | | | No DIF |
| | | | What city are we in | N/A | | | No DIF |
| | | DIF among tentative linking items, treating confident items as anchors | | | | | |
| | | | Floor of building | N/A | | | No DIF |
| | | | Address | N/A | | | No DIF |
| Mex-Cog | Orientation | | | | | | |
| | | DIF among confident linking items | | | | | |
| | | | Day of month | N/A | | | No DIF |
| | | | Month | N/A | | | No DIF |
| | | | Year | N/A | | | No DIF |
| | | | Day of the week | N/A | | | No DIF |
| | | | What state are we in | N/A | | | No DIF |
| | | DIF among tentative linking items, treating confident items as anchors | | | | | |
| | | | None | | | | |
| CHARLS HCAP | Orientation | | | | | | |
| | | DIF among confident linking items | | | | | |
| | | | Year | 0.71 | 0.66 | 0.77 | Negligible |
| | | | Day of the week | 0.44 | 0.41 | 0.48 | DIF |
| | | | What state are we in | 0.66 | 0.57 | 0.76 | Negligible |
| | | | What county are we in | 1.50 | 1.38 | 1.62 | Negligible |
| | | | Address | 1.79 | 1.63 | 1.96 | DIF |
| | | | Day of month | N/A | | | No DIF |

| | | |
|---|---|---|
| Month | N/A | No DIF |
| What city are we in | N/A | No DIF |
| Floor of building | N/A | No DIF |
| DIF among tentative linking items, treating confident items as anchors | | |
| Season of year | N/A | No DIF |

Supplemental Figure 1: Flowchart of item banking procedure implemented to statistically harmonize cognition across HCAP studies

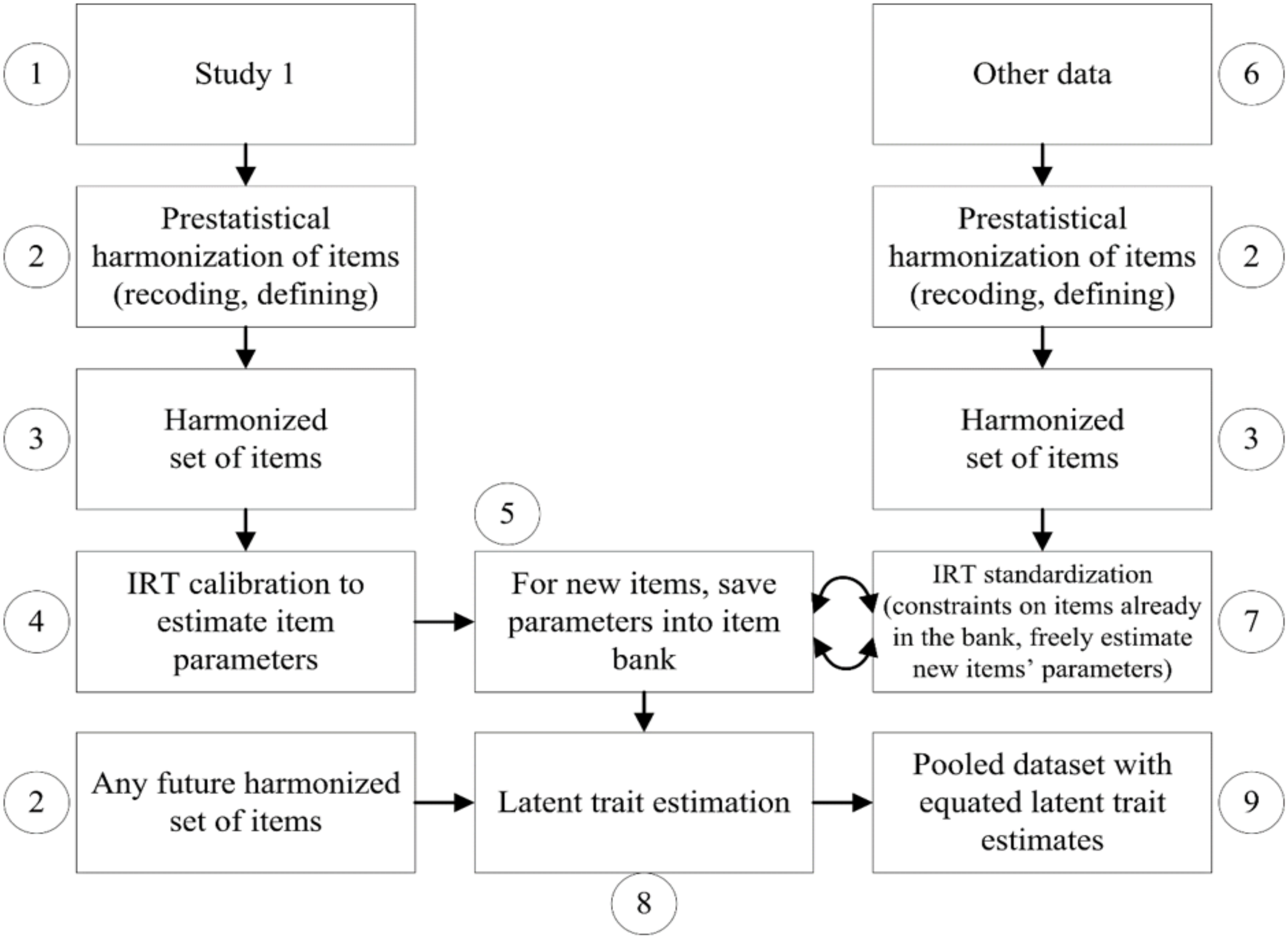

Legend. This procedure was implemented separately for cognitive domains of orientation, memory, executive functioning, language, and general cognitive performance. Starting with a reference study, HRS-HCAP (step 1), pre-statistical harmonization was conducted by recoding and redefining cognitive test items as needed. Minor transformations were performed as needed to handle missing data and outlying values (step 2), resulting in a harmonized set of cognitive test items to support a measurement model (step 3). Step 4 entails calibration via item response theory methods (equivalent to confirmatory factor analysis, CFA) to freely estimate item parameters including factor loadings, and thresholds (for categorical test items) or intercepts (for continuous test items). Resulting item parameters are saved into an item bank (step 5). Next, additional studies are serially brought in (step 6) to have their cognitive test items recoded as necessary in the same manner as in other studies (step 2 at right), resulting in a unique set of harmonized cognitive test items for a study (step 3 at right). In step 7, IRT standardization is implemented with a CFA model that places constraints on parameters for items in common between the other study and HRS-HCAP as well as previous studies already processed, and freely estimates parameters for items unique to the new study. Parameters for these new items are iteratively added to the item bank (step 5), such that eventually any future harmonized set of items can be used to estimate latent traits (step 8) and save out factor scores into a pooled dataset for all included studies (step 9).
